# Seasonal variation in SARS-CoV-2 transmission in the Netherlands, 2020-2022: statistical evidence for an inverse association with solar radiation and temperature

**DOI:** 10.1101/2024.11.28.24318154

**Authors:** Don Klinkenberg, Jantien Backer, Chantal Reusken, Jacco Wallinga

## Abstract

In temperate regions, respiratory viruses such as SARS-CoV-2 are better transmitted in winter than in summer. Understanding how the weather is associated with SARS-CoV-2 transmissibility can enhance projections of COVID-19 incidence and improve estimation of the effectiveness of control measures. During the pandemic, transmissibility was tracked by the reproduction number *R*_*t*_. This study aims to determine whether information about the daily temperature, absolute humidity, and solar radiation improves predictions of *R*_*t*_ in the Netherlands from 2020 to 2022, and to quantify the relationship between *R*_*t*_ and daily weather data. We conducted a regression analysis, accounting for immunity from vaccination and previous infection, higher transmissibility of new variants, and changes in contact behaviour due to control measures. Results show a linear association between log*R*_*t*_ and daily solar radiation and temperature, indicating a ratio of *R*_*t*_ in Winter versus Summer of 1.7 (95% CI, 1.4-2.1). The possibility that this association arises from unrelated seasonal patterns was dismissed, as weather data from earlier years provided poorer fits with only small effect sizes. This suggests a causal relationship of solar radiation and temperature with SARS-CoV-2 transmissibility, enhancing confidence in using this relationship for short-term predictions and other epidemiological analyses.

## BACKGROUND

Many infectious diseases display seasonal patterns in incidence caused by seasonality in exposure (e.g. Lyme disease, Q-fever) or seasonality in contact rates (e.g. measles),[1] as well as by other mechanisms. With many pathogens, associations of transmission potential with weather variables have been found, mainly temperature and humidity.[1-3] Associations with weather variables are especially pronounced for respiratory viruses, where the causal mechanism may be related to the pathogen (e.g. virus stability related to temperature, or better persistence in drier air related to aerosole size),[4-6] the individual host (amount and duration of shedding, or antiviral defense)[4, 6] or host-host interaction (e.g. people spending less time indoors during warm and sunny weather, where most transmission takes place).[5, 7, 8]

Already early in the COVID-19 pandemic a relation between weather variables and SARS-CoV-2 transmission has been hypothesized and tested, to assess the risk for a large second wave by the end of 2020 in northern temperate regions.[9-11] Many studies focused on the transmission potential, which can be expressed as the time-dependent reproduction number *R*_*t*_, defined as the mean number of secondary cases infected per primary case. Large-scale studies across many geographical locations showed associations of *R*_*t*_ with temperature and humidity, with often small and non-linear effect sizes. A study among 409 cities in 26 countries worldwide found a maximum *R*_*t*_ at a temperature around 10°C and at an absolute humidity around 6.6 g/m^3^, with effect sizes of 0.087 and 0.061 compared to minimal values at 20°C and 11 g/m^3^, respectively.[12] In a multi-county analysis in mainland USA, almost 20% of variation in *R*_*t*_ was explained by temperature, humidity and ultraviolet radiation, with low *R*_*t*_ values when the weather is hot, humid, and sunny. Overall, the effects were concentrated at the extreme ends of the ranges of the variables. For instance, *R*_*t*_ was largest at a temperature of 20°C, by about 5% compared to any other temperate under 30°C; and specific humidity affected transmissibility only at very low or very high values, suggesting that the model better explained spatial than seasonal variation in *R*_*t*_.[13] In a study in 47 prefectures in Japan, the reproduction number *R* was found to decrease with about 6% when comparing temperatures of 0°C vs 30°C, and with 5% when absolute humidity increased to 10g/m^3^ or higher.[14] In general, across many studies, temperature was found to be inversely associated with SARS-CoV-2 transmission, whereas the association with humidity was unclear.[15] In Europe, SARS-CoV-2 transmission was most often found to be inversely related to temperature, humidity, and solar radiation.[5]

The impact of weather on SARS-CoV-2 transmission has significant implications for epidemiological analyses, such as forecasting incidence by dynamic transmission models or estimating the effects of sets of control measures. To account for a weather effect in such analyses, it should be properly and independently quantified for the target population at hand. This requires the target population to be sufficiently large, so that the epidemic is driven by transmission events within the population, affected by the local weather, instead of reflecting epidemics in other locations by many import cases. It also requires the epidemic in the target population to have a sufficiently uniform climate, because the mechanisms by which weather may impact transmissibility are not self-evident, and may be different in different climates. The importance of focussing on single target populations is further illustrated by the fact that estimated effect sizes for smaller populations tend to be large, e.g. 46% higher *R*_*t*_ in winter compared to summer explained by temperature and absolute humidity in France,[16] or 22% in London and Paris,[17] whereas those from larger populations are smaller or reflect spatial rather than seasonal variation.[13, 14]

In this study, we estimated the association between SARS-CoV-2 transmissibility and temperature, absolute humidity, and solar radiation during 25 months of the pandemic in the Netherlands. We quantified this relationship and estimated the relative change in reproduction number in winter vs summer resulting from the association. We assessed whether the estimated association can be attributed to the general seasonal trend of temperature, absolute humidity, and solar radiation, or to the specific course of the weather during the pandemic.

## METHODS

We used the log of the daily *instantaneous* reproduction number *R*_*t*_, which was estimated during the pandemic by the National Institute of Public Health and the Environment (RIVM) in the Netherlands (Figure 1a),[18, 19] and the variance of the estimator. It was estimated by dividing the symptom onset incidence at time *t* + 5 (incubation period is estimated to be 5 days) by the number of infectious individuals, i.e. the symptom onset incidence in the days up to *t* + 5 weighted by the serial interval distribution.[20] Because of autocorrelation and day-of-week effects in the time series of daily estimated *R*_*t*_, we only used the time series of log*R*_*t*_ estimates of Fridays (Figure 1a, see supplementary material S1). The observation period was from 22 February 2020 until 29 March 2022, after which *R*_*t*_ estimates became less reliable due to changes in case notification policy.

**Figure 1.**
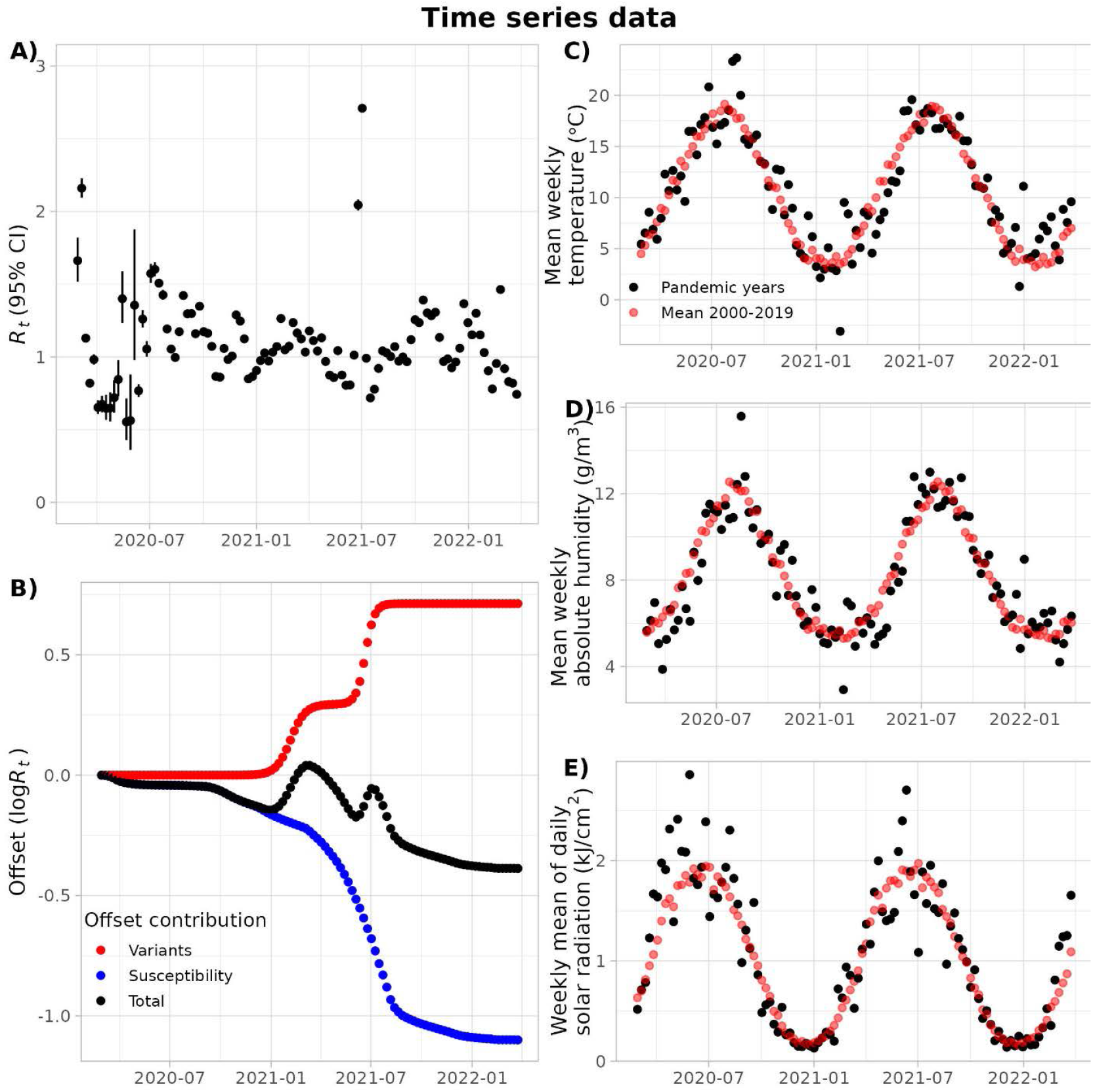
The time course of all datasets. (A) the response variable log*R*_*t*_, with the 95% confidence interval indicating the uncertainty of the estimator, the inverse of which is used to weigh the observations; (B) the offsets due to increased transmissibility from new variants, and decreased transmissibility from immunity due to infection and vaccination; (C) the mean weekly temperature during the pandemic and in the years 2000-2019; (D) the mean weekly absolute humidity during the pandemic and in the years 2000-2019; (E) the weekly mean of daily solar radiation during the pandemic and in the years 2000-2019.

Weekly genomic surveillance data were obtained from RIVM’s open data.[21] They were used to estimate 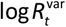, i.e. the relative mean transmissibility at time *t*, caused by the circulating variants Alpha, Beta, Gamma, and Delta, relative to the wildtype variant (Figure 1b, see supplementary material S2).[22] After the emergence of Delta, 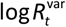 was kept constant, because the emergence of Omicron in December 2021 was for an important part due to immune escape.

Cumulative incidence CumIncid(*t*) per day of the pandemic was calculated by RIVM, by combining serological data and daily hospitalisation data, and previously used to calculate the prevalence of infected individuals.[23] It was available until 2 July 2021, after which CumIncid(*t*) was kept constant, because waning immunity and re-infections with new variants made estimation unreliable. Vaccine coverage data were obtained from RIVM’s open data.[24] We used the weekly national coverage of the completed primary vaccination, interpolated to obtain daily coverages. To calculate the rise of vaccine-induced immunity VaccImm(*t*) in the population, the coverage was multiplied by 75% vaccine effectiveness as estimated against the Delta variant,[25] because the primary vaccination series was quickly followed by the rise of the Delta variant. Cumulative incidence and vaccine-induced immunity were combined to calculate the times series of the susceptible fraction of the population as *s*_*t*_ = (1 - CumIncid(*t*)) (1 - VaccImm(*t*)) (Figure 1b). Note that *s*_*t*_ only reflects the decrease in susceptibilty until the Summer of 2021, after which waning immunity and the rise of the Omicron variant made reliable estimation impossible.

The time series (109 weeks) of weekly reproduction numbers from 22 February 2020 to 29 March 2022 was partitioned into 41 intervention periods (see supplementary material S3), separated by changes in COVID-19 control measures or the start or end of school holidays. After a preliminary analysis for outlier detection based on Cook’s distance (supplementary material S1),[26] the period between 26 June 2021 and 9 July 2021 was split into two separate weeks for the main analyses. This was a period when bars and nightclubs were allowed to open, causing a short sudden increase in *R*_*t*_ (Figure 1a) followed by a rapid surge in new cases and closure of bars and nightclubs in the period thereafter.

We used the weekly averaged (Tuesday to Monday) temperature *T*_*t*_ (degrees Celsius), absolute humidity *ah*_*t*_ (g/m^3^), and solar radiation *sr*_*t*_ (kJ/cm^2^) as weather variables (Figure 1c,d). We used observations from the “De Bilt” weather station, located centrally in the Netherlands,[27] and calculated absolute humidity from relative humidity and temperature as

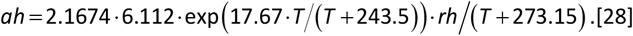

We used the Oxford Stringency Index for the Netherlands as a compound measure for the stringency of control measures (Oxford COVID-19 Government Response Tracker).[29] It was used to indicate changes in control measures and compare these to changes in the estimated regression coefficients.

We conducted weighted time series regression analyses to estimate the association of the SARS-CoV-2 reproduction number *R*_*t*_ and weather variables between 22 February 2020 and 29 March 2022 in the Netherlands. The full regression equation for the main analysis was

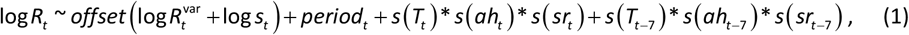

in which the lags are 7 days because of the use of weekly data, and weekly averaged weather variables. The observations were weighted by 1 var(log *R*_*t*_) to reflect uncertainty in the estimated log*R*_*t*_. Offsets were included to account for a known increase in transmissibility because of the emergence of new variants (up to and including Delta), and for a known decrease in the susceptible fraction resulting from immunity due to natural infection (until 2 July 2021) and primary vaccination. The factor *period* was added to correct for the effect of different contact rates due to changing intervention measures or school holidays and to absorb other mechanisms for gradual changes in transmissibility, such as compliance to measures, behaviour unrelated to measures, and population immunity not included in the offsets, by booster vaccination, natural waning of immunity and immune escape due to the emergence of the various Omicron variants.

We fitted 57 models with the last six terms of equation (1), i.e. temperature, absolute humidity, and/or solar radiation, with or without interaction terms, in the same week or lagged by one week, with a linear relation or with thin-plate regression splines. For this, we used generalised additive models (R package mgcv,[30] version 1.9.0), with the REML optimisation method. We compared the models by AIC_c_, Akaike Information Criterion with an additional correction[31] because of the high number of parameters (including 41 periods) relative to observations (109 weeks): *AIC*_*c*_ = *AIC* + 2× *df* ×(*df* + 1) (*n* - *df* - 1), in which AIC and *df* are calculated output from the mgcv package (*df* is estimated with spline models). A lower AIC_c_ by at least 2 points difference indicates a better model. All AIC_c_ values are reported as ΔAIC_c_, relative to the model in equation (1) without the weather terms. We translated estimated model parameters to a mean *R*_*t*_ in winter vs summer, by first calculating 7-day rolling averages of temperature, absolute humidity, and solar radiation per day-of-year with the mean weather of 2000-2019, then predicting a daily *R*_*t*_, and finally dividing the maximum weekly *R*_*t*_ by the minimum.

We challenged our results against the possibility that they are due to day-of-year as a confounder, i.e. that there was an association between log*R*_*t*_ and the weather only because both are seasonal. We did this by fitting models with mean daily temperature, absolute humidity, and solar radiation of the years 2000-2019, and with the five time series with the exact weather data of earlier years, all starting with a leap year: 2000-2002, 2004-2006, 2008-2010, 2012-2014, and 2016-2018. This addressed the question if log*R*_*t*_ can be explained by the general seasonal trend of the weather rather than the specific fluctuations in temperature,absolute humidity, and solar radiation in the two years of the pandemic.

We did sensitivity analyses to address some specific choices we made. First, we repeated the main analysis by using averages of weather variables across 28 weather stations in the Netherlands, weighted by local population size. Second, we repeated the main analysis with estimates of reproduction numbers for different days than Friday. Third, we repeated the main analysis with a single two-week period from 26 June 2021 to 9 July 2021, instead of two separate one-week periods. Fourth, we repeated the main analyses without offsets, to assess if the *period* variable will compensate for this; if so, we can be more confident that *period* will also absorb other gradually changing effects, in particular loss of immunity and the rise of the Omicron variant.

## RESULTS

We see a clear preference (lower AIC_c_) for two linear models, both with an inverse association with solar radiation: one with temperature and the other with absolute humidity (Table 1). Splines and delay terms never improved model fits (lower AIC_*c*_); including interaction terms improved model fits only for models with both temperature and absolute humidity (supplementary material S4).

**Table 1.** Supports and estimated seasonal effects of models in the main analysis, including offsets and *period*. The selected model with lowest ΔAIC_c_ is bold-faced.

| | Model coefficients (SE) <sup>a</sup> | | | | $R_t$ winter vs summer<br>(95% CI) | Performance | |
| --- | --- | --- | --- | --- | --- | --- | --- |
| | Temperature | Absolute humidity | Solar insolation | Interaction T and ah <sup>b</sup> | | $\Delta AIC_c$ <sup>c</sup> | $R^2$ <sup>d</sup> |
| 0 | - | - | - | - | 1 | 0 | 0.68 |
| 1 | -0.023 (0.006) | - | - | - | 1.5 (1.2 ; 1.7) | -19.2 | 0.74 |
| 2 | - | -0.049 (0.015) | - | - | 1.5 (1.2 ; 1.8) | -10.5 | 0.72 |
| 3 |  |  | -0.16 (0.05) | - | 1.3 (1.1 ; 1.6) | -8.3 | 0.71 |
| 4 | -0.032 (0.013) | 0.027 (0.034) | - | - | 1.4 (1.2 ; 1.7) | -14.6 | 0.74 |
| 5 | -0.060 (0.016) | 0.059 (0.034) | - | -0.0046 (0.0017) | 1.8 (1.4 ; 2.4) | -20.6 | 0.76 |
| 6 | **-0.020 (0.005)** | - | **-0.12 (0.05)** | - | **1.7 (1.4 ; 2.1)** | **-23.3** | **0.76** |
| 7 | - | **-0.051 (0.014)** | **-0.16 (0.05)** | - | **1.9 (1.5 ; 2.4)** | **-22.4** | **0.75** |
| 8 | -0.013 (0.015) | -0.019 (0.038) | -0.14 (0.06) | - | 1.8 (1.4 ; 2.3) | -17.9 | 0.75 |
| 9 | -0.041 (0.020) | 0.021 (0.042) | -0.090 (0.060) | -0.0036 (0.0018) | 2.0 (1.5 ; 2.7) | -18.4 | 0.76 |

<sup>a</sup> Measurement units: temperature (°C), absolute humidity (g/m<sup>3</sup>), solar radiation (kJ/cm<sup>2</sup>); <sup>b</sup> Interaction between temperature and absolute humidity; other interaction terms never improved model fit as measured by $AIC_c$ ; <sup>c</sup> Corrected Akaike's Information Criterion, relative to that of the least complex model (first line); <sup>d</sup> R-squared, proportion of variance explained

According to the model with temperature and solar radiation, *R*_*t*_ is expected to decrease by 2.0% (standard error 0.5%) with every degree Celsius increase, and by 12% (s.e. 5%) with every kJ/cm^2^ increase of solar radiation, which results in a ratio of *R*_*t*_ in winter (around 1 February) vs summer (around 1 August) of 1.7 (95% CI: 1.4; 2.1). The model with absolute humidity and solar radiation predicts this ratio to be 1.9 (1.5; 2.4) (Table 1, Figure 2).

**Figure 2.**
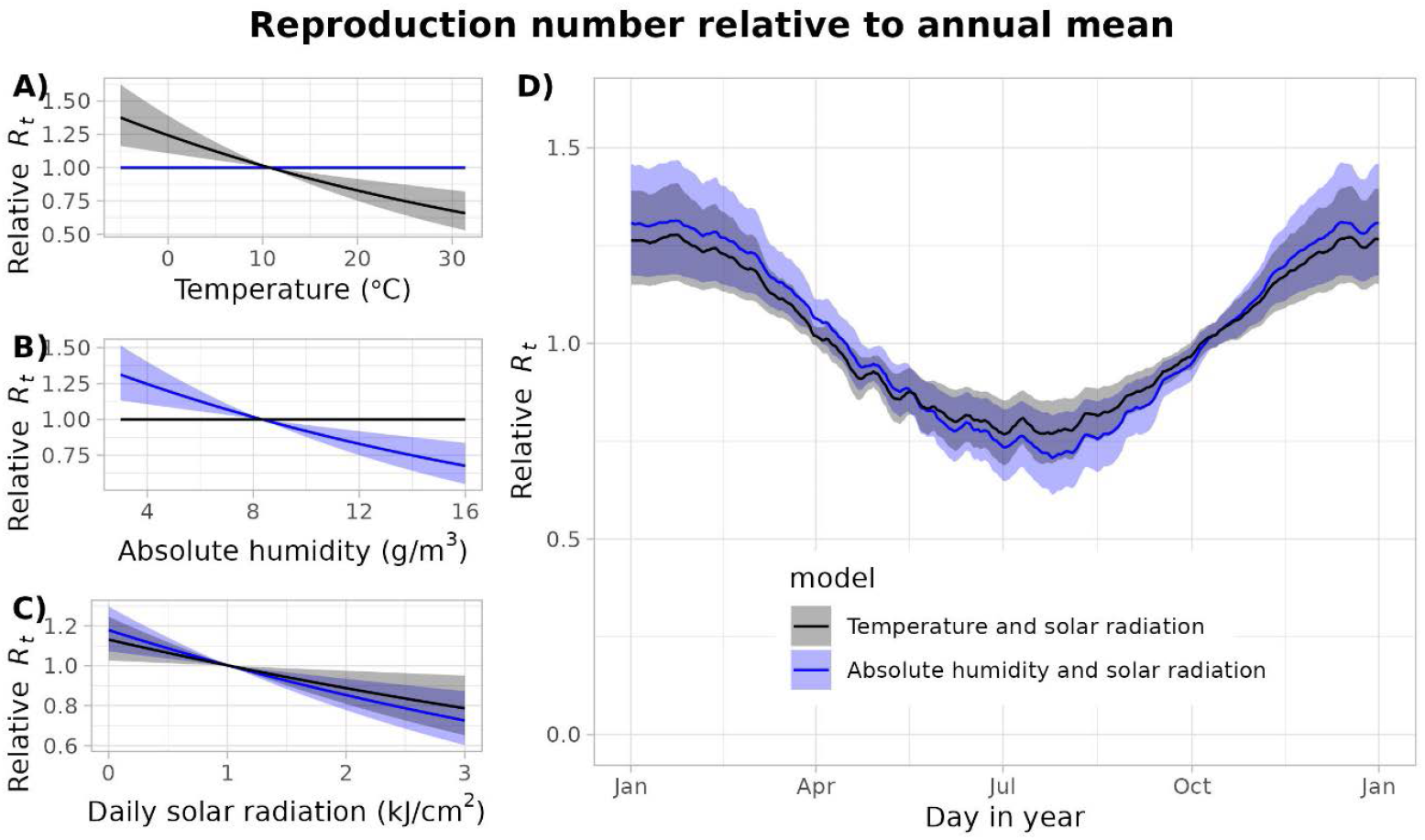
The change in reproduction number due to the associations with temperature and solar radiation, and with absolute humidity and solar radiation, expressed relative to the annual mean. (A) The direct relation with temperature; (B) the direct relation with absolute humidity; (C) the direct relation with daily solar radiation; (C) the annual cycle using the relation between reproduction number and the mean daily weather variables of the years 2000-2019.

*Period* and offsets explain 68% of the variance in log*R*_*t*_ (Table 1). The estimates of the period effects turn out to reflect the stringency of control measures very well, as illustrated by the qualitative agreement with the Oxford Stringency Index (Figures 3, S2): if measures become more stringent, the reproduction number decreases. This qualitative agreement ends in Autumn 2021, after which the period-associated reproduction number tends to be higher, especially when Omicron arises in 2022.

**Figure 3.**
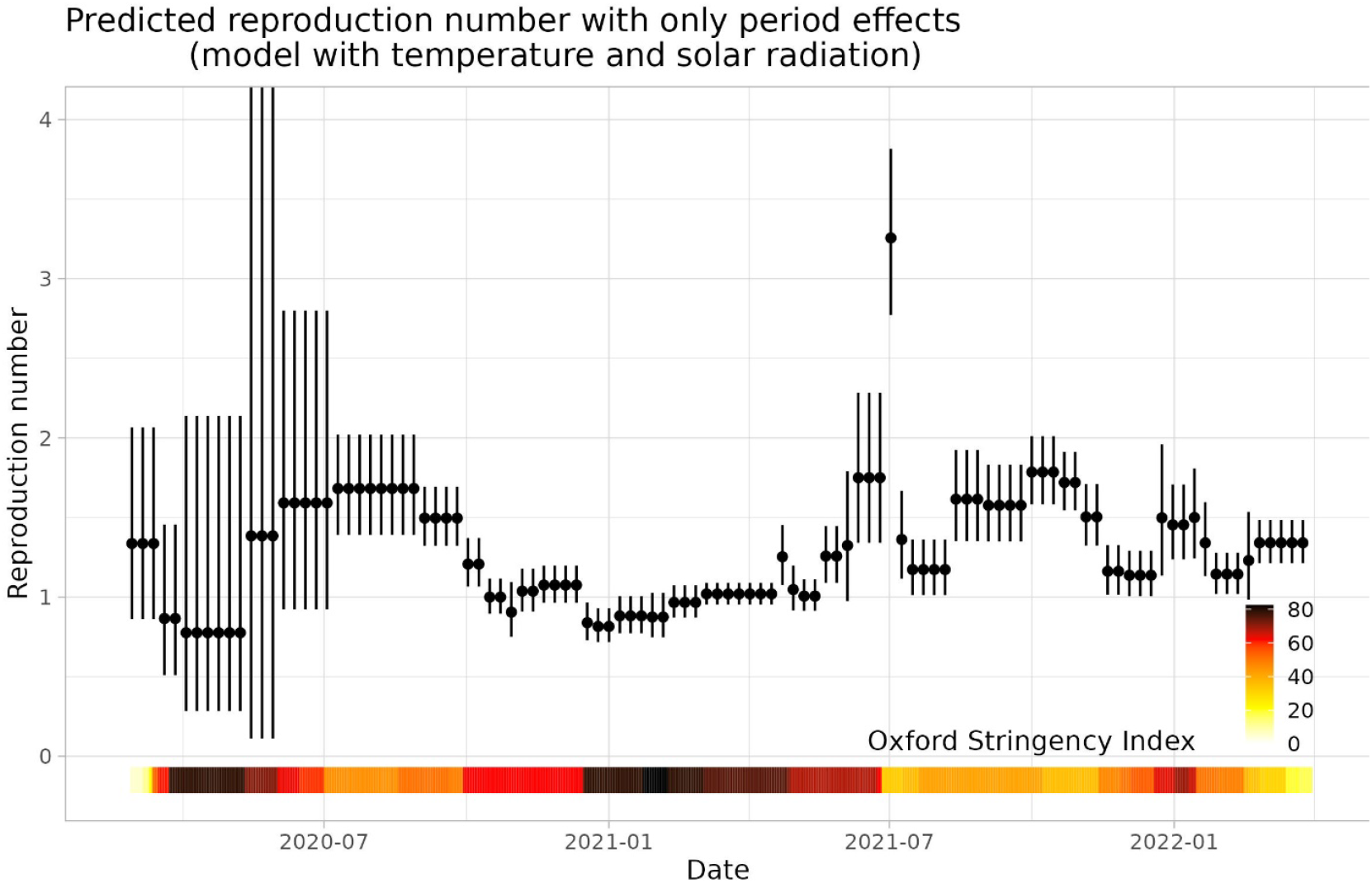
The reproduction number during the pandemic with mean annual weather and without offsets, as predicted by the model with linear temperature and solar radiation effects. It shows the contribution of the *period* estimates, mainly reflecting the effectiveness of interventions and adherence to interventions, but after the summer of 2021 also waning of immunity and the emergence of the Omicron variants. The coloured bar indicates the Oxford Stringency index, a compound measure (scale 0-100) of overall stringency of government interventions[29].

The effect of solar radiation and temperature (or absolute humidity) in determining overall transmissibility explained another 8% (or 7%) of variance in log*R*_*t*_ (Table 1). This is clearly visible when comparing the observed *R*_*t*_ and the predictions from the models with and without weather variables (Figures 4, S3): especially during longer periods without changes in control measures (June to October 2020, and February to April 2021), fluctuations in *R*_*t*_ are much better explained in the model with weather variables.

**Figure 4.**
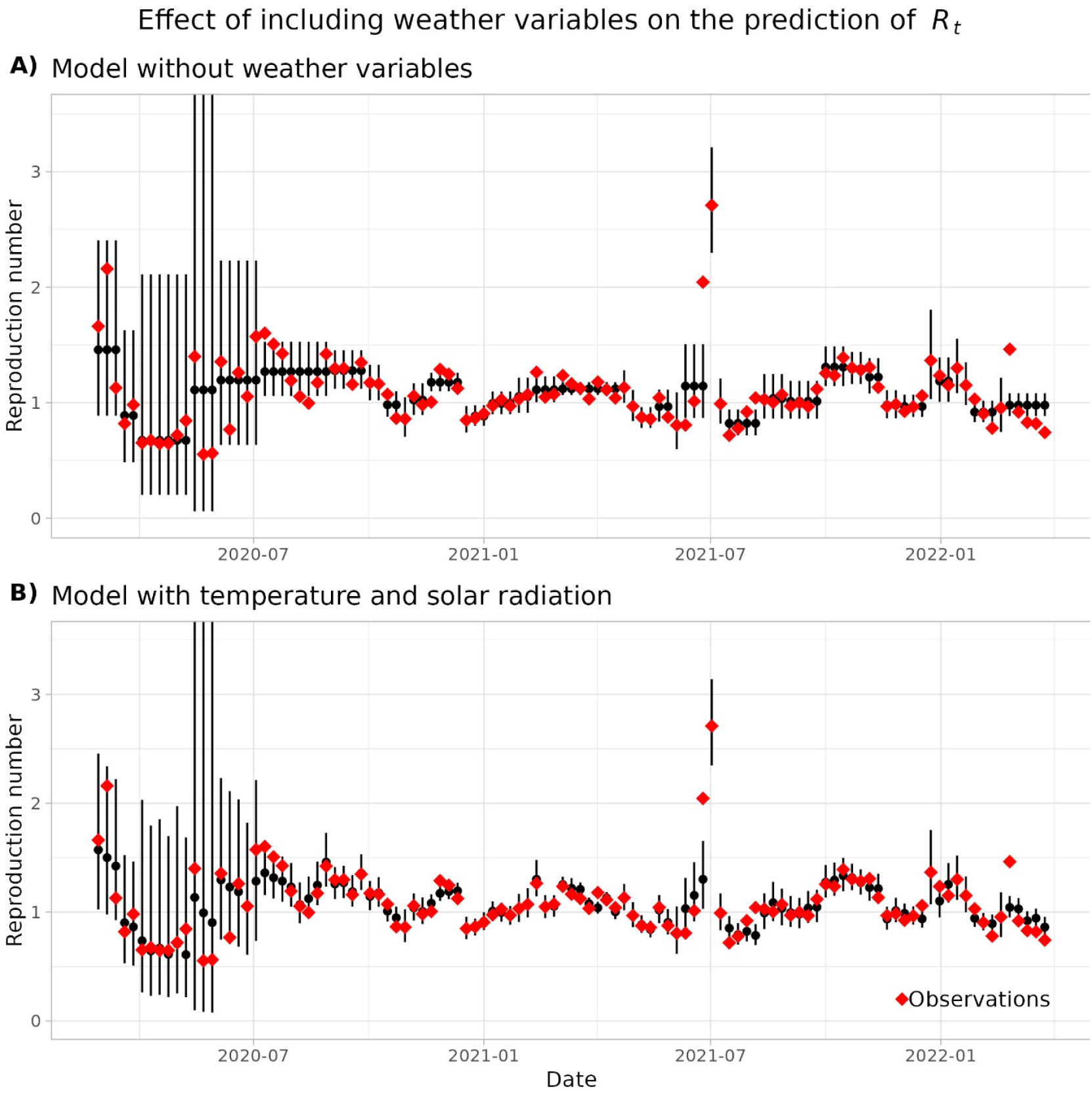
The reproduction number during the pandemic, predicted by (A) the model with only offsets and *period*, and (B) the model with linear temperature and solar radiation effects.

To assess robustness of these results, we addressed the question if the association of *R*_*t*_ with weather variables was confounded by day of the year, i.e. if the association is only there because both are seasonal. We did this by fitting the log*R*_*t*_ data to alternative time series of temperature, absolute humidity, and/or solar radiation: the means of the years 2000-2019, and five periods of exact measurements in earlier years, with all models presented in the main results. There were only eight combinations of weather variables and models showing better fits than the model without weather variables (most with data from 2016-2018), suggesting some association to a general seasonal pattern, but none of the 54 models explained log*R*_*t*_ better than the exact weather data from 2020-2022 (Table 2). The estimated effect sizes (*R*_*t*_ in winter vs summer) do indicate a small general seasonal effect explained by the weather variables, but never as large and significant as with the original 2020-2022 data (Figure 5).

**Table 2.**
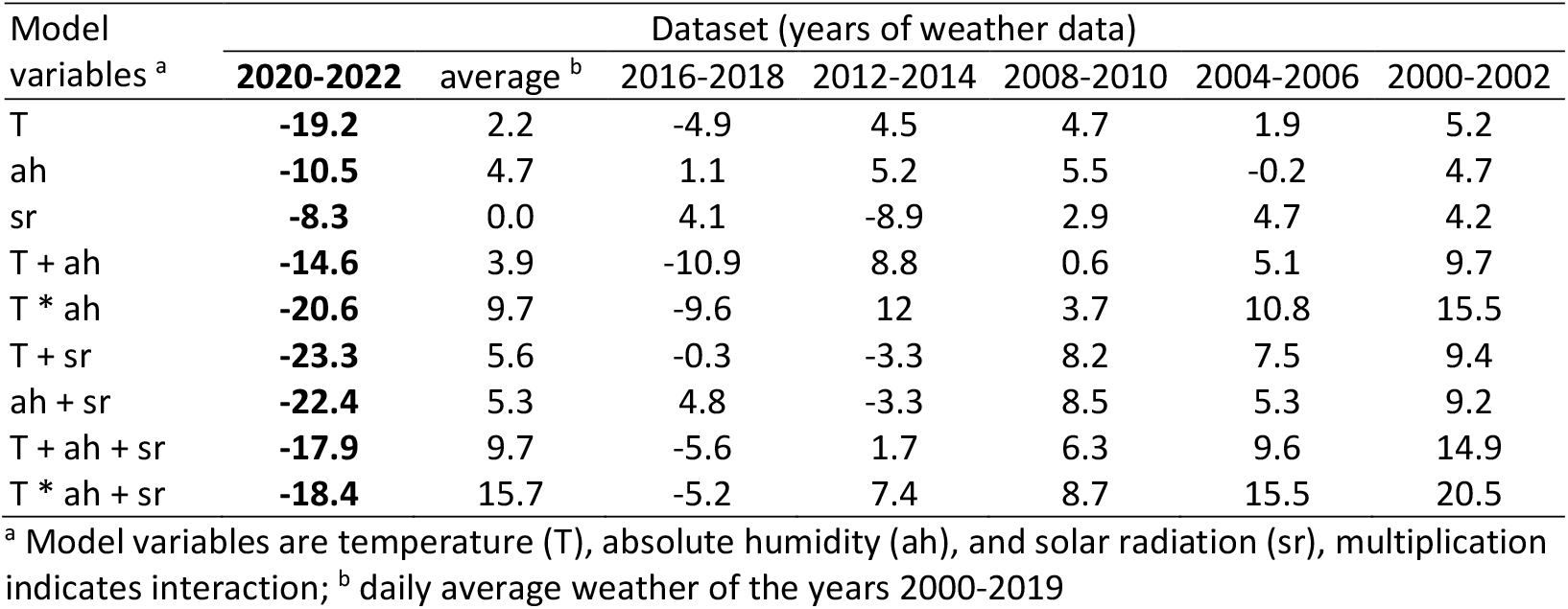
Additional analysis adressing confounding of weather variables and the reproduction number *R*_*t*_ by day-of-year (co-seasonality). The table shows ΔAIC_c_ values of model fits with the original weather time series during the pandemic and alternative time series of other years (lowest in bold). ΔAIC_c_ is calculated relative to that of the least complex model, i.e. without weather variables.

| Model variables <sup>a</sup> | Dataset (years of weather data) |  |  |  |  |  |  |
| --- | --- | --- | --- | --- | --- | --- | --- |
|  | **2020-2022** | average <sup>b</sup> | 2016-2018 | 2012-2014 | 2008-2010 | 2004-2006 | 2000-2002 |
| T | **-19.2** | 2.2 | -4.9 | 4.5 | 4.7 | 1.9 | 5.2 |
| ah | **-10.5** | 4.7 | 1.1 | 5.2 | 5.5 | -0.2 | 4.7 |
| sr | **-8.3** | 0.0 | 4.1 | -8.9 | 2.9 | 4.7 | 4.2 |
| T + ah | **-14.6** | 3.9 | -10.9 | 8.8 | 0.6 | 5.1 | 9.7 |
| T * ah | **-20.6** | 9.7 | -9.6 | 12 | 3.7 | 10.8 | 15.5 |
| T + sr | **-23.3** | 5.6 | -0.3 | -3.3 | 8.2 | 7.5 | 9.4 |
| ah + sr | **-22.4** | 5.3 | 4.8 | -3.3 | 8.5 | 5.3 | 9.2 |
| T + ah + sr | **-17.9** | 9.7 | -5.6 | 1.7 | 6.3 | 9.6 | 14.9 |
| T * ah + sr | **-18.4** | 15.7 | -5.2 | 7.4 | 8.7 | 15.5 | 20.5 |

<sup>a</sup> Model variables are temperature (T), absolute humidity (ah), and solar radiation (sr), multiplication indicates interaction; <sup>b</sup> daily average weather of the years 2000-2019

**Figure 5.**
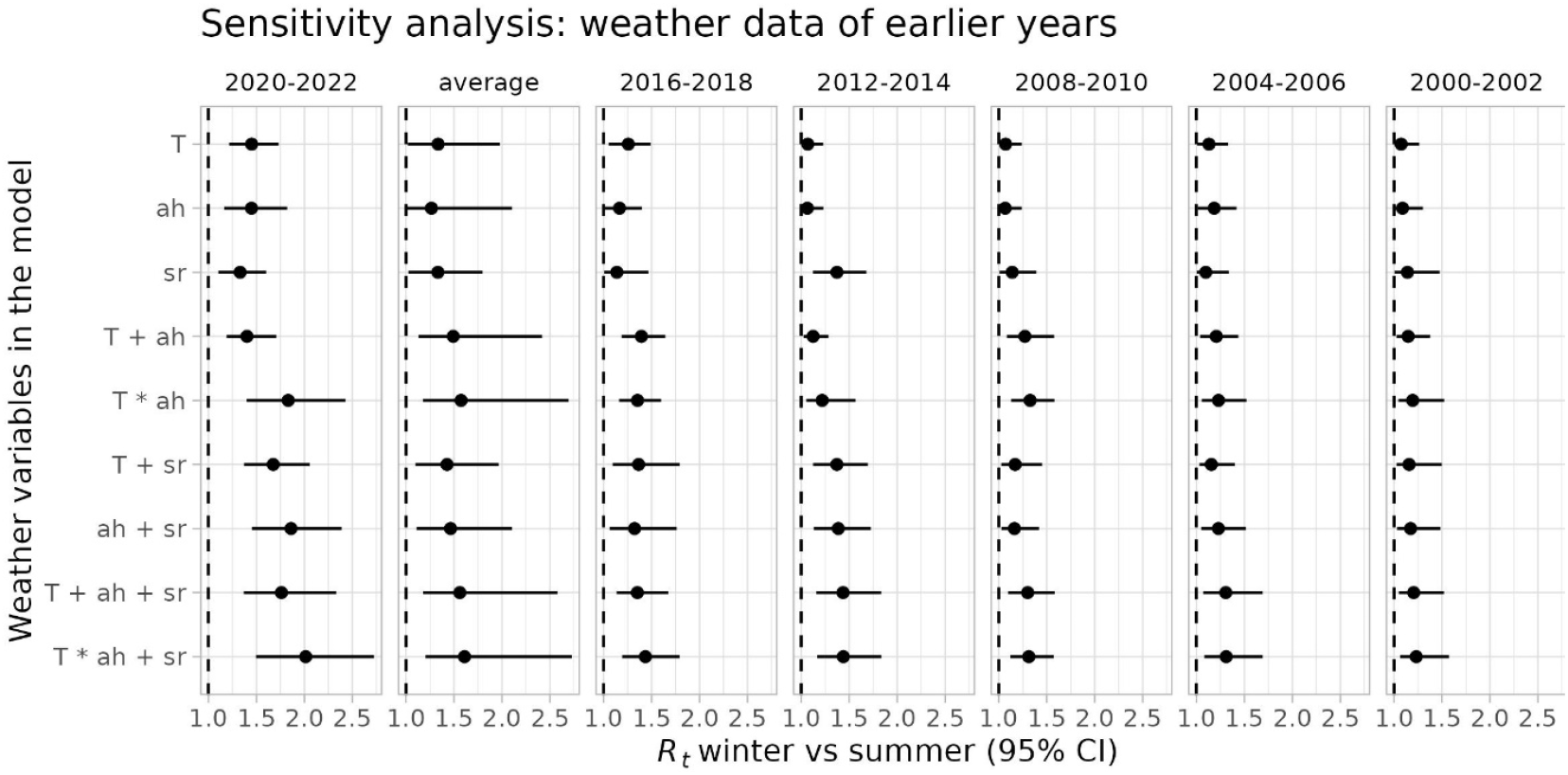
Additional analysis adressing confounding of weather variables and the reproduction number *R*_*t*_ by day-of-year (co-seasonality). The figure shows the magnitude of the estimated seasonal effect (*R*_*t*_ in winter vs summer, 95% CI) associated with the original weather time series during the pandemic, the average weather between 2000-2019, and alternative time series of other years.Weather variables in the models are temperature (T), absolute humidity (ah), and solar radiation (sr); multiplication indicates inclusion of an interaction term.

Sensitivity analyses show that our decision to use the data from a single weather station did not affect our results (supplementary material S5). We repeated our main analyses with log*R*_*t*_ time series of other days than Fridays (supplementary material S5), and found that our final model with temperature and solar radiation as linear terms had highest or considerable support (AIC_c_ within two points of the minimum) with data from all days except Wednesday and Thursday. Across all weekday datasets, the model with only temperature and no solar radiation was also supported very well. The datasets of Tuesdays and especially Wednesdays showed lowest support overall for associations of *R*_*t*_ with the weather, which supports our decision for using the Friday data, because the Tuesday and Wednesday data were estimated with incidences of Sundays and Mondays, which were most affected by day-of-the-week effects in reporting. The estimated ratio of reproduction number *R*_*t*_ in winter vs summer varied between 1.2 and 1.9, with the estimates of the main analysis at the higher end of this range.

We also repeated the main analysis with a single two-week period between 26 June 2021 and 9 July 2021 (supplementary material S5), which resulted in more complex models with interactions receiving most support: the model with temperature and solar radiation with an estimated *R*_*t*_ in winter vs summer of 2.5, and the model absolute humidity and solar radiation with an estimated *R*_*t*_ in Winter vs Summer of 2.9. This finding confirms that results were sensitive to the two data points indicated by evaluating Cook’s distance in the preliminary analysis, thus justifying our choice of periods to use two separate one-week periods and reduce the influence of these data points.

Finally, we repeated the main analysis without the offset term, to check if the *period* variable indeed absorbs gradual changes in *R*_*t*_ due to effects not included in the model. Leaving out the offset did not change the estimated effects of the weather variables, but did indeed result in *period* effects incorporating the offsets (supplementary material S5).

## DISCUSSION

We estimated the association between transmissibility of SARS-CoV-2 during the pandemic in the Netherlands, and the mean daily temperature, absolute humidity, and solar radiation. We found an inverse linear association with temperature and solar radiation to explain 8% of variance in log*R*_*t*_ during the pandemic, with reproduction number *R*_*t*_ decreasing by 2.0% for every degree Celsius increase, and by 12% for every kJ/cm^2^ daily solar energy. Extrapolating to mean weekly temperatures during the year, this means that *R*_*t*_ early February (Winter) is 70% higher than *R*_*t*_ early August (Summer) in the Netherlands. A model receiving similar support suggests an association with absolute humidity and solar radiation, translated to a seasonal effect of a 90% higher *R*_*t*_ in winter vs summer.

To estimate the effect of weather variables, we had to separate this effect from the much larger effect on SARS-CoV transmission during the pandemic by several other factors. The effects of variants, immunity from infection, and vaccination could be included as offsets up to Autumn 2021, after which waning immunity and immune escape due to the Omicron variant obscured the level of immunity in the population. However, the varying sets of sometimes rapidly changing social distancing measures had even greater impacts which could not be estimated independently. To account for these, the stepwise constant *period* was added to the model with changepoints at the times of changing control measures and school holidays. These could explain large changes in log*R*_*t*_ (68% of variance) due to changing control policies, but also allowed for more gradual changes due to other effects such as changing behaviour due to epidemic fatigue, and changes in immunity or variant transmissibility not accounted for in the offsets. The fact that the estimated effect of the weather variables does not change when leaving out the offsets (supplementary material S5) shows that such gradual changes are indeed absorbed by the *period* variable. Including the offsets is useful however, because it allows us to interpret the coefficients for *period* as the effects of control measures, which could therefore be compared with the Oxford Stringency Index as a qualitative validation (Figure 2a).[29]

We addressed the possibility that the association was only a result of two seasonally co-oscillating variables without any direct or indirectly causal link, by fitting log*R*_*t*_ to alternative datasets of temperature, absolute humidity, and solar radiation in the Netherlands on the same dates but in different years, and a 20-year average. A small seasonal effect was observed with all datasets (Figure 5), but with most datasets and models (46/54) there was no statistical evidence for an association between log*R*_*t*_ and weather variables at all, and with each model log*R*_*t*_ was best explained by the data from 2020-2022, the years of the pandemic. This result is a strong indication that the actual weather was causally related with SARS-CoV-2 transmission during the pandemic. Regardless of the causal nature of the relationship, changes in the actual solar radiation and temperature (or absolute humidity) are thus good predictors of changes in transmissibility. Prediction models can take advantage of this by using actual weather data to anticipate changes in incidence that will result from such changes in transmissibility.

A strength of our study was that we had a high-quality *R*_*t*_ time series of more than two years. However, the *R*_*t*_ estimates were autocorrelated and *R*_*t*_ was highly affected by changing behaviour due to control measures and holidays, but also related to other factors such as pandemic.[32] Changes in *R*_*t*_ due to variants and immunity by infection or vaccination could be accounted for, but only up to the summer of 2021, after which waning immunity and the rise of the Omicron variant made population immunity untrackable. We accounted for these issues by using only weekly data and including a *period* variable to avoid assumptions on changing behaviour and immunity. However, this resulted in a considerable loss of degrees of freedom and therefore statistical power. We used AIC_*c*_ for model selection, which strongly favoured simpler model with linear terms rather than splines, without delays, and with no or limited interactions, whereas many earlier studies have shown complex relations between *R*_*t*_ and temperature, humidity and solar or UV radiation. The limitation in statistical power also made it difficult to correct for more possibly relevant variables, such as mobility data, which we considered but resulted in unstable models due to high collinearity with especially solar radiation (results not shown).

Earlier studies on associations between weather variables and SARS-CoV-2 transmission generally agreed on an inverse effect of temperature. Results on the role of humidity were less consistent. Our study shows evidence of an inverse association with solar radiation, in combination with temperature or absolute humidity, resulting in a larger seasonal amplitude than estimated before in similar climates.[16, 17] Because most SARS-CoV-2 transmission took place indoors,[8] it is likely that this association is for a large part due to people going outside in warm and sunny weather. However, even then the true mechanism is still unknown: people may have fewer or shorter close contacts outside, but it does not rule out the possibility of host or virus factors, e.g. sunlight affecting virus stability. The fact that our results are inconclusive about the role of temperature or absolute humidity may have to do with the limited statistical power for more complex models, as discussed above. Given that the existing literature is more conclusive about temperature,[15] and that our sensitivity analyses with datasets from other weekdays generally favour models with temperature over those with absolute humidity (supplementary material S5), we have more confidence in a role for temperature rather than absolute humidity.

Unique to our study is that we aimed to quantify changes in SARS-CoV-2 transmissibility attributable to changes in weather variables, to assess the magnitude of seasonal variation in transmissibility in the Netherlands. For a reliable quantification it is important to restrict estimation to a limited geographic area, because the relationship may differ in different geographic locations, making a single estimate from multiple locations difficult to interpret. Keeping the model simple also adds to reliability of the estimation, because if weather variables are included with many other explanatory variables, there is a risk that the different variables are dependent or display spurious associations, resulting in potentially biased estimates. Our results suggest that using solar radiation and temperature as covariates will increase the performance of prediction models of SARS-CoV-2 transmissibility and resulting COVID-19 incidence.

## CONCLUSIONS

In the Netherlands during the COVID-19 pandemic, SARS-CoV-2 transmissibility was higher with less solar radiation and with lower temperature. This resulted in a 70% higher reproduction number in winter compared to summer. Sensitivity analyses confirmed a robust association with weather variables, suggesting a valuable role of weather data data for prediction models of COVID-19 incidence.

## Data Availability

All data and code in the present work are available online at github.com/rivm-syso/covid-seasonality

## ACKNOWLEDGEMENTS

We thank all collaborators in the Data Analytics, Research, and Automated Reporting (DARA) team in the Center for Infectious Disease Control for processing and curating all COVID-19 surveillance data. We thank Dr Scott McDonald for valuable discussions on time series analyses.

No additional funding was received for this study.

Data and codeall:data and code are available online at github.com/rivm-syso/covid-seasonality

