## Supplemental Online Appendices for "Seasonal variation in SARS-CoV-2 transmission in the Netherlands, 2020-2022: statistical evidence for an inverse association with solar radiation and temperature"

Contents

Section 1: Preliminary analyses: autocorrelation and identification of outliers
Section 2: Relative infectivity due to new variants (offset $\log R_t^{\text{var}}$ )
Section 3: Intervention periods
Section 4: Additional results
Section 5: Sensitivity analyses

Section 1: Preliminary analyses: autocorrelation and identification of outliers

*Autocorrelation*

The analyses were done with weekly $\log R_t$ estimates for SARS-CoV-2 in the Netherlands instead of daily estimates, to prevent autocorrelation between the observations. Daily estimates were expected to be autocorrelated because the denominator of the estimator

$$\hat{R}_t = \frac{x_{t+5}}{\sum_{\tau} g_{\tau} x_{t+5-\tau}} \qquad \text{(S1)}$$

is highly correlated for subsequent days. Here, $x_t$ is the incidence of reported cases by day of symptom onset and $g_\tau$ is the serial interval distribution[1,2]; we shift the estimates of $R_t$ back into the past by the mean incubation period of 5 days, such that time $t$ corresponds to the time of infection. We expected the numerator to cause day-of-week effects because testing behaviour differed between days of the week, causing irregularities in reported symptom onsets because of recall biases (especially Mondays were overrepresented at the cost of Sundays). We decided to use $\log R_t$ of Fridays, because the numerator of that estimator is number of reported symptom onsets on Wednesdays, minimizing the risk of irregularities in reporting due to the weekend.

To support the decision to use weekly data, we fitted eight models in total, with general structure

$$\log R_t \sim offsets + period\left[+day\_of\_week\right]\left[+\sum \log R_{t-\tau}\right]\left[+T_t + ah_t + sr_t\right] \qquad \text{(S2)}$$

Models differed by:

- using daily data or weekly data (Fridays only). When using daily data, *day_of_week* was added to account for day-of-week effects in reporting
- adding lagged $\log(R_t)$ terms or not. With daily data (time measured in days), lagged terms were added for a delay of up to a week ($t-7 \le \tau \le t-1$). With weekly data (time measured in weeks), only $\log(R_{t-7})$ was added
- adding weather variables or not, i.e. temperature, absolute humidity, and solar radiation with linear terms. When using daily data, the observations of single days were used. With weekly data, weekly averages were used (from the Tuesday before to the Monday after each Friday)

We tested in all analyses for the presence of first-order autocorrelation with the Breusch-Godfrey test (in the package lmtest[3] in R statistical software, Table S1) and visualized autocorrelation in ACF-plots (Figure S1). The models with daily data and without lagged terms had residual autocorrelation, shown by the test ($p < 0.05$) and ACF-plot. Adding lagged terms resolved significance of the autocorrelation, but the ACF plot seems to indicate some remaining negative first-order autocorrelation. With weekly data, there was no residual autocorrelation.

We did a sensitivity analysis with other days of the week to address our decision to use Friday data (Supplementary Data, section 4).

Table S1. Breusch-Godfrey *p*-values for eight models, run with either daily or weekly data, with or without lagged $\log R_t$ terms, and with or without weather variables (both temperature and absolute humidity), testing for the presence of first-order residual autocorrelation

| Model | Daily or weekly | Delay terms | Weather variables | Breusch-Godfrey *P* |
|---|---|---|---|---|
| 1 | Daily | | | 0.000 |
| 2 | Daily | | Yes | 0.000 |
| 3 | Daily | Yes | | 0.120 |
| 4 | Daily | Yes | Yes | 0.003 |
| 5 | Weekly | | | 0.783 |
| 6 | Weekly | | Yes | 0.218 |
| 7 | Weekly | Yes | | 0.805 |
| 8 | Weekly | Yes | Yes | 0.908 |

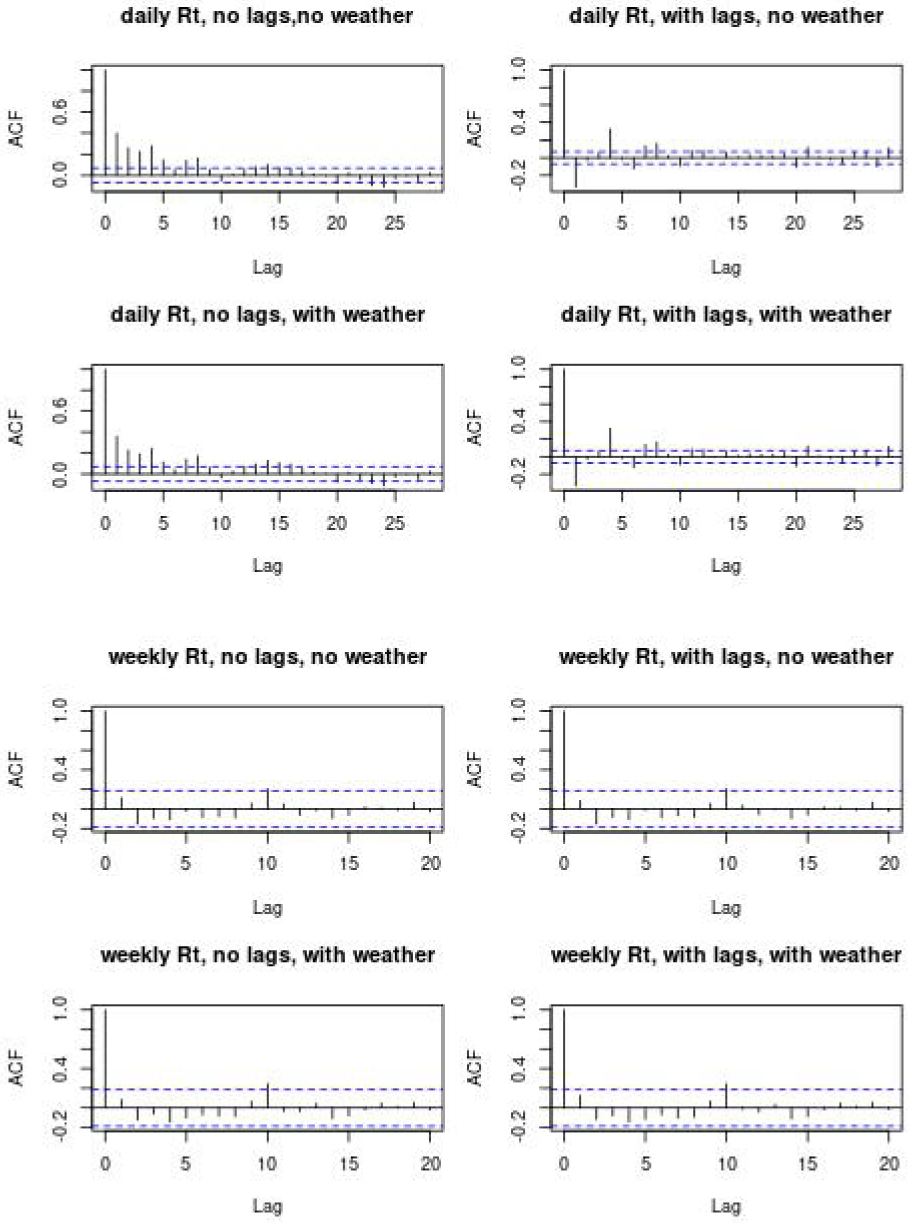

Figure S1. Autocorrelation function plots of eight models, run with either daily or weekly data, with or without lagged $\log R_t$ terms, and with or without weather variables (temperature, absolute humidity, and solar radiation). Presence of autocorrelation at a particular lag is indicated by the vertical bar at that lag (X-axis) exceeding the dotted horizontal line.

*Influential datapoints*

We split the two-week period from 26 June 2021 to 9 July 2021 into two separate periods of one week, because the two datapoints in this period were highly influential. That decision was based on a preliminary fit of four models with general structure of equation (S2), without *day_of_week* or lagged terms. Models differed by:

- either a single two-week period from 26 June 2021 to 9 July 2021, or two one-week periods
- inclusion of weather terms, i.e. temperature, absolute humidity, and solar radiation (linear terms)

We calculated Cook's distances[4] of all datapoints (Figure S2). The two datapoints of the two-week period were very influential when combined into a single period, with Cook's Distance > 1 for one of the points, which is generally considered too high. The problem was resolved by splitting the period into two one-week periods.

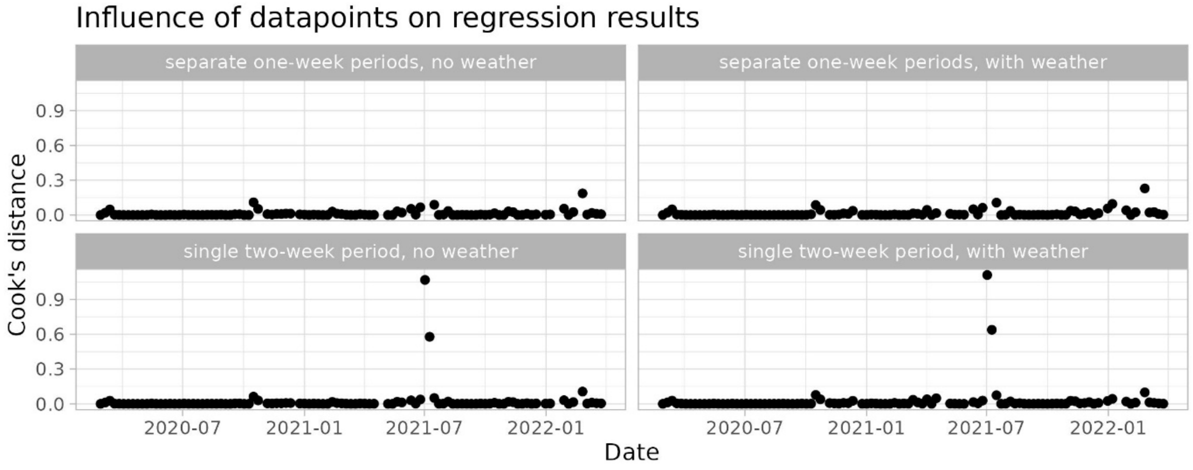


Figure S2. Cook's distances of all datapoints, showing two highly influential datapoints when using a single period from 26 June 2021 to 9 July 2021.

Section 2: Relative infectivity due to new variants (offset $\log R_t^{\text{var}}$)

The relative infectivity due to new variants at time *t* is calculated as

$$R_t^{\text{var}} = \sum_v prev_{t,v} \beta_v \tag{S3}$$

in which $prev_{t,v}$ is the relative prevalence of infectious individuals of variant *v* at time *t*, and $\beta_v$ is the relative transmissibility of variant *v* compared to the wildtype virus *v* = 1. Both $prev_{t,v}$ and $\beta_v$ are estimated from genomic surveillance data as follows.

The genomic surveillance data[5] report the genomic variants of SARS-CoV-2 isolates from randomly sampled patients per week in the Netherlands. We removed all samples that are not wildtype, Alpha variant, Beta variant, Gamma variant, and Delta variant, and used data up to and including 10 October 2021, when all samples were Delta variant. We fitted a multinomial logistic regression model to the data, with time *t* as dependent variable[6]. This resulted in estimates of the relative growth rates $\lambda_v$ of variant *v* relative to the wildtype virus *v* = 1, and it resulted in fitted curves for the proportions of variants reported, $pr_{t,v}^{rep}$. To obtain the proportions of variants in daily incidence of infections, we shift the curve 3.5 days to the past, combining a shift of 3.5 days to the future because the weeks in the data are indicated by their first day (Monday), and shift of 7 days to the past because of 5 days incubation plus 2 days interval between symptom onset and sampling: $pr_{t,v}^{\inf} = pr_{t+3.5,v}^{rep}$.

From the estimated relative growth rates $\lambda_v$, we calculated the relative infectivities $\beta_v$ of all variants relative to the wildtype. The relation between growth rate $\lambda$ and infectivity $\beta$ is derived by using the SEEIIR transmission model we used to create a generation interval distribution that matched observations early in the pandemic[7]:

$$\begin{aligned}
\frac{ds}{dt} &= -\beta s\left(i^{(1)} + i^{(2)}\right) \\
\frac{de^{(1)}}{dt} &= \beta s\left(i^{(1)} + i^{(2)}\right) - \gamma e^{(1)} \\
\frac{de^{(2)}}{dt} &= \gamma\left(e^{(1)} - e^{(2)}\right) \\
\frac{di^{(1)}}{dt} &= \gamma\left(e^{(2)} - i^{(1)}\right) \\
\frac{di^{(2)}}{dt} &= \gamma\left(i^{(1)} - i^{(2)}\right) \\
\frac{dr}{dt} &= \gamma i^{(2)}
\end{aligned} \quad . \tag{S4}$$

In this system, the variables are the proportions of individuals in the population in the susceptible (*s*), the two exposed ($e^{(1)}$ and $e^{(2)}$), the two infected-and-infectious ($i^{(1)}$ and $i^{(2)}$), and recovered (*r*) classes. The parameter $\gamma$ determines the rate of progression through the states of infection, and is equal to 0.875 per day to obtain a mean generation interval of 4 days. The system can be solved by assuming exponential growth with rate $\lambda$, so that

- Ansatz: $e^{(1)}(t)=e_1 \exp(\lambda t)$ and $e^{(2)}(t)=e_2 \exp(\lambda t)$
- Using Eq (S4), part 3:
  $\lambda e_2 \exp(\lambda t)=\gamma(e_1-e_2)\exp(\lambda t) \Rightarrow$
  $e_2=e_1 \cdot \frac{\gamma}{\gamma+\lambda} \Rightarrow$
  $e^{(2)}(t)=e_1 \cdot \frac{\gamma}{\gamma+\lambda} \exp(\lambda t)$
- Continuing with parts 4 and 5:
  $i^{(1)}(t)=e_1\left(\frac{\gamma}{\gamma+\lambda}\right)^2 \exp(\lambda t)$ and
  $i^{(2)}(t)=e_1\left(\frac{\gamma}{\gamma+\lambda}\right)^3 \exp(\lambda t)$
- Filling these into part 2:
  $\lambda e_1 \exp(\lambda t)=\beta s e_1\left(\left(\frac{\gamma}{\gamma+\lambda}\right)^2+\left(\frac{\gamma}{\gamma+\lambda}\right)^3\right)\exp(\lambda t)-\gamma e_1 \exp(\lambda t)$, resulting in

$$\beta=\frac{1}{s} \cdot \frac{(\gamma+\lambda)^4}{2\gamma^3+\gamma^2\lambda}. \qquad \text{(S5)}$$

When we assume that the wildtype epidemic was at a constant level when the Alpha variant emerged, i.e. $\lambda_1$ = 0, the relative transmissibility of each variant relative to the wildtype can be calculated as

$$\beta_v=\left[\frac{1}{s} \cdot \frac{(\gamma+\lambda_v)^4}{2\gamma^3+\gamma^2\lambda_v}\right] \Big/ \left[\frac{1}{s} \cdot \frac{\gamma^4}{2\gamma^3}\right]=\frac{2(\gamma+\lambda_v)^4}{2\gamma^4+\gamma^3\lambda_v}. \qquad \text{(S6)}$$

As a last step, we calculate the relative prevalences $prev_{v,t}$ of the variants $v$ from the relative incidences $pr_{v,t}^{\inf}$ and relative transmissibilities $\beta_v$, by using the incidence of infection term in equation (S4), part 1:

$$\begin{aligned} pr_{v,t}^{\inf} &\propto \beta_v s \cdot prev_{v,t} \Rightarrow \\ prev_{v,t} &= \frac{pr_{v,t}^{\inf} / \beta_v}{\sum_j pr_{j,t}^{\inf} / \beta_j}, \end{aligned} \qquad \text{(S7)}$$

where index $j$ runs over the circulating variants.

Section 3: intervention periods

During the COVID-19 pandemic we used a simulation model to make short-term projections of hospital and ICU admissions[7]. In that model, during the pandemic, we defined change points on dates at which control measures changed or when school holidays started or ended. We used these change points to define the *period* variable. A very brief description of the periods, with the day they ended:

2020-03-13: no control
2020-03-28: first lockdown
2020-05-10: first lockdown, second phase (Klinkenberg et al, 2024)
2020-06-01: primary schools and businesses partly open
2020-07-05: primary and secondary schools open, more leisure allowed
2020-08-30: more leisure allowed, Summer holiday
2020-09-28: schools fully open, less work from home
2020-10-14: work from home, limited group sizes, bars close early
2020-10-25: smaller group sizes, bars closed, school holiday
2020-11-04: no school holiday
2020-11-18: two weeks of more restricted leisure and sport
2020-12-14: back to period before
2020-12-20: schools closed, businesses closed, all leisure closed
2021-01-03: Christmas holiday
2021-01-22: back to period before
2021-02-07: curfew after 9pm
2021-02-28: primary schools open
2021-04-18: secondary schools partly open, some restricted shopping
2021-04-27: child care open
2021-05-02: curfew lifted, universities partly open, businesses open, bars outside open
2021-05-16: May holiday
2021-05-18: end of May Holiday
2021-05-30: more leisure, inside sports
2021-06-04: secondary schools fully open
2021-06-25: larger group sizes, more leisure, restaurants open
2021-07-09: wide relaxation including nightlife open, Summer holiday
2021-08-08: nightlife closed, Summer holiday
2021-08-31: no change [extra period break]
2021-09-24: end Summer holiday
2021-10-17: further relaxations in bars, theaters
2021-10-31: school holiday
2021-11-12: back to period before
2021-11-27: early closure bars, restaurants, businesses
2021-12-18: further restrictions in opening hours
2021-12-24: back to lockdown, including school closures
2022-01-09: Christmas holiday
2022-01-16: schools open
2022-01-25: universities open, businesses partly open
2022-02-17: bars open, most leisure allowed
2022-02-24: businesses fully open
2022-04-01: remaining restrictions lifted

Section 4: additional results

Table S2. $AIC_c$ values and estimates of $R_t$ in winter vs summer (95% CI) of all models in the main analysis. For each set of weather variables (inside a box), the simplest model with lowest $AIC_c$ is bold-faced

| Weather variables in model | Linear | | Splines | |
|---|---|---|---|---|
| | Without delays | With delays | Without delays | With delays |
| - | 0 | 0 | 0 | 0 |
| T | **-19.2** | -13.6 | -15.1 | -9.2 |
| | **1.4 (1.2 ; 1.7)** | 1.4 (1.1 ; 1.8) | 1.8 (1.4 ; 2.3) | 1.8 (1.3 ; 2.4) |
| ah | **-10.5** | -6.2 | -10.5 | -6.2 |
| | **1.4 (1.2 ; 1.8)** | 1.3 (1.1 ; 1.8) | 1.4 (1.2 ; 1.8) | 1.3 (1.1 ; 1.8) |
| si | -8.3 | -11.6 | -6.2 | -6.2 |
| | 1.3 (1.1 ; 1.6) | 1.6 (1.3 ; 2.0) | 1.3 (1.1 ; 1.6) | 1.9 (1.5 ; 2.6) |
| T + ah | -14.6 | -6.9 | -9.4 | 2.5 |
| | 1.4 (1.2 ; 1.7) | 1.4 (1.2 ; 1.7) | 1.7 (1.3 ; 2.3) | 1.8 (1.4 ; 2.5) |
| T * ah | **-20.6** | -9.0 | -5.5 | 24.5 |
| | **1.8 (1.4 ; 2.4)** | 1.8 (1.3 ; 2.5) | 1.7 (1.3 ; 2.3) | 1.9 (1.4 ; 2.7) |
| T + si | **-23.3** | -16.6 | -18.4 | -2.7 |
| | **1.7 (1.4 ; 2.1)** | 1.8 (1.4 ; 2.3) | 1.9 (1.5 ; 2.5) | 2.1 (1.6 ; 2.9) |
| T * si | -21.1 | -10.5 | -11.3 | 4.8 |
| | 1.8 (1.4 ; 2.3) | 2.0 (1.5 ; 2.8) | 1.9 (1.5 ; 2.5) | 2.1 (1.5 ; 2.9) |
| ah + si | **-22.4** | -16.6 | -22.1 | -5.5 |
| | **1.9 (1.4 ; 2.4)** | 1.8 (1.4 ; 2.5) | 1.9 (1.4 ; 2.4) | 2.0 (1.5 ; 2.7) |
| ah * si | -19.2 | -11.0 | -22.4 | -16.6 |
| | 2.1 (1.5 ; 2.8) | 2.1 (1.5 ; 3.1) | 1.9 (1.5 ; 2.4) | (1.8 ; 2.5) |
| T + ah + si | -17.9 | -4.7 | -12.0 | 17.9 |
| | 1.8 (1.4 ; 2.3) | 1.8 (1.3 ; 2.6) | 2.0 (1.5 ; 2.6) | 2.1 (1.5 ; 3.1) |
| T * ah + si | **-18.4** | 1.3 | -5.9 | 27.1 |
| | **2.0 (1.5 ; 2.7)** | 2.0 (1.4 ; 2.9) | 1.9 (1.4 ; 2.6) | 2.1 (1.5 ; 3.1) |
| T * si + ah | -15.7 | 2.0 | -5.0 | 24.0 |
| | 1.9 (1.5 ; 2.6) | 2.1 (1.4 ; 3.1) | 1.9 (1.5 ; 2.6) | 2.1 (1.5 ; 3.1) |
| T + ah * si | -15.0 | 1.8 | -12.2 | 5.3 |
| | 2.0 (1.5 ; 2.7) | 2.1 (1.5 ; 3.1) | 2.0 (1.5 ; 2.6) | 2.0 (1.4 ; 2.9) |
| T * ah * si | -0.4 | 40.8 | [a] | [a] |
| | 2.0 (1.4 ; 3.0) | 2.1 (1.4 ; 3.3) | | |

[a] too complex model

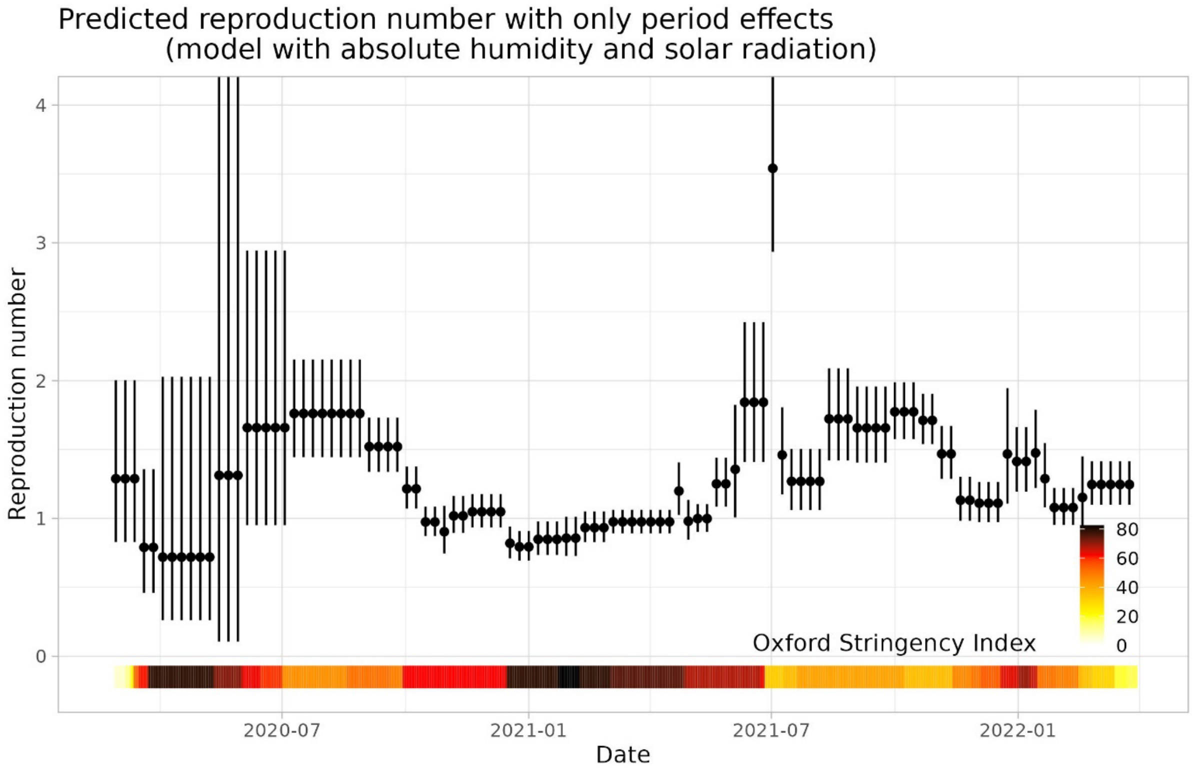


Figure S2. The reproduction number during the pandemic with mean annual weather and without offsets, as predicted by the model with linear absolute humidity and solar radiation effects. It shows the contribution of the *period* estimates, mainly reflecting the effectiveness of interventions and adherence to interventions, but after the summer of 2021 also waning of immunity and the emergence of the Omicron variants. The coloured bar indicates the Oxford Stringency index, a compound measure (scale 0-100) of overall stringency of government interventions[8].

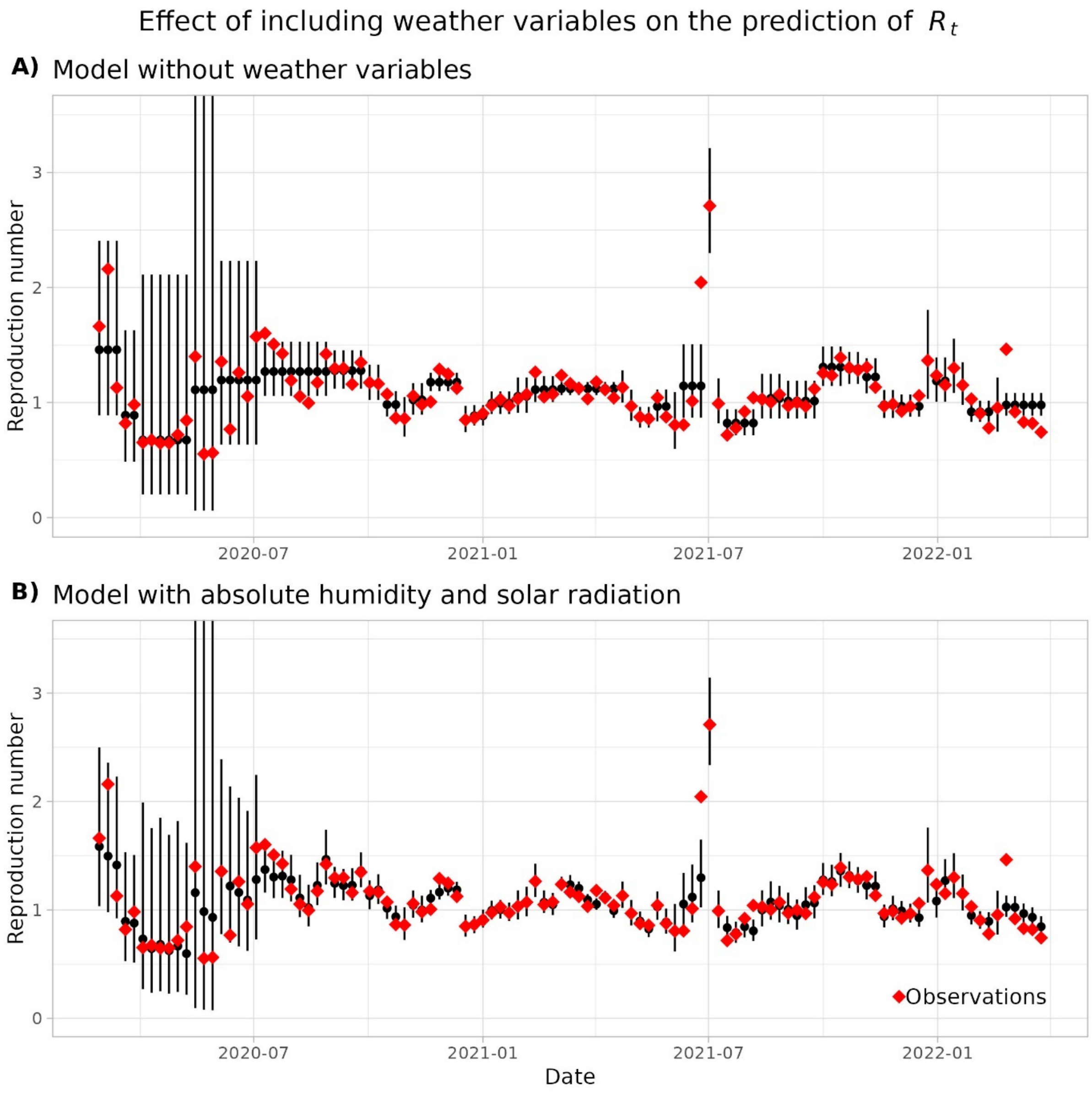

Figure S3. The reproduction number during the pandemic, predicted by (A) the model with only offsets and *period*, and (B) the model with linear absolute humidity and solar radiation effects.

Section 5: sensitivity analyses

*Dataset with weather data averaged across weather stations*

The main analysis was done with the weather data from the "De Bilt" weather station, located centrally in The Netherlands, whereas the response variable log$R_t$ was a national average. For this sensitivity analysis, we used an average temperature, absolute humidity and solar radiation across 28 weather stations, weighted by local population size.

To weigh the weather stations in the weather data (ref knmi), we used the information on https://www.weerverlet.nl/knmi-weerstation/ (visited 18 June 2025), which links weather stations to postcodes to inform weather conditions on construction sites. Population sizes on postcode level were obtained from Statistics Netherlands (www.cbs.nl).

Table S3 is the Results Table for these data, presented as Table 1 from the main text. Differences from the main results are very small, with the same two models having most support.

Table S3. Results as in the main analysis, but with weather data averaged across the Netherlands, showing model coefficients, estimated seasonal effects, and model performance.

| | Model coefficients (SE) [a] | | | | $R_t$ winter vs summer (95% CI) | Performance | |
|---|---|---|---|---|---|---|---|
| | Temperature | Absolute humidity | Solar radiation | Interaction T and ah [b] | | $\Delta AIC_c$ [c] | $R^2$ [d] |
| 0 | - | - | - | - | 1 | 0 | 0.68 |
| 1 | -0.022 (0.006) | - | - | - | 1.4 (1.2 ; 1.7) | -17.7 | 0.73 |
| 2 | - | -0.050 (0.015) | - | - | 1.5 (1.2 ; 1.8) | -11.2 | 0.72 |
| 3 | | | -0.16 (0.05) | - | 1.3 (1.1 ; 1.6) | -7.9 | 0.71 |
| 4 | -0.026 (0.013) | 0.012 (0.034) | - | - | 1.4 (1.2 ; 1.8) | -12.2 | 0.73 |
| 5 | -0.052 (0.016) | 0.041 (0.035) | - | -0.0043 (0.0017) | 1.8 (1.4 ; 2.5) | -16.6 | 0.75 |
| 6 | -0.020 (0.006) | - | -0.12 (0.05) | - | 1.7 (1.3 ; 2.0) | -21.0 | 0.75 |
| 7 | - | -0.052 (0.014) | -0.16 (0.05) | - | 1.9 (1.4 ; 2.4) | -22.4 | 0.75 |
| 8 | -0.006 (0.015) | -0.038 (0.038) | -0.15 (0.06) | - | 1.8 (1.4 ; 2.4) | -16.8 | 0.75 |
| 9 | -0.030 (0.021) | -0.003 (0.043) | -0.11 (0.06) | -0.0030 (0.0018) | 2.0 (1.5 ; 2.8) | -15.2 | 0.76 |

[a] Measurement units: temperature (C), absolute humidity (g/m$^3$), solar radiation (kJ/cm$^2$); [b] Other interaction terms never improved model fit as measured by $AIC_c$; [c] Corrected Akaike's Information Criterion, relative to that of the least complex model (first line); [d] R-squared, proportion of variance explained

*Datasets with log$R_t$ from other weekdays*

The main analysis was done with log$R_t$ estimates of Fridays, because these were estimated from symptom onset incidence on Wednesdays, which was least disturbed by irregularities due to weekend effects. Here we check how robust our results are to this choice.

Tables S2 and S3 show the $\Delta AIC_c$ and estimated effects of the weather on $R_t$ (Winter vs Summer) of all models of the main analysis, estimated with log$R_t$ from different days of the week. With all datasets, the best model (lowest $AIC_c$) of our main analysis had the lowest $AIC_c$ as well, or was within 2 points of the lowest $AIC_c$ indicating similar support. The estimated effect of temperature on $R_t$ with the linear model (1.5 with Friday data) was lower with the data from Monday – Thursday, in particular Tuesday and Wednesday. However, the best spline models with the Wednesday and

Thursday data resulted in larger estimates. In conclusion, the results in the main text are robust to the choice of weekday for log$R_t$.

Table S4. $\Delta$AIC$_c$ values of main analyses with datasets of different weekdays. Lowest values per weekday are underlined; values within 2 points are in dark grey cells; within 5 in light grey.

| Model terms | | | Weekday | | | | | | |
|---|---|---|---|---|---|---|---|---|---|
| *T* | *ah* | *sr* | Fri | Sat | Sun | Mon | Tue | Wed | Thu |
| Linear models | | | | | | | | | |
| - | - | - | 0 | 0 | 0 | 0 | 0 | 0 | 0 |
| X | - | - | -19.2 | -21.0 | -29.2 | **-27.4** | **-10.8** | -5.7 | -14.6 |
| - | X | - | -10.5 | -10.4 | -16.7 | -16.1 | -1.5 | 0.8 | -9.7 |
| - | - | X | -8.3 | -14.0 | -17.4 | -7.2 | -0.7 | 0.6 | 1.2 |
| X | X | - | -14.6 | -17.5 | -24.7 | -22.6 | -10.5 | -4.3 | -9.1 |
| X | - | X | **-23.3** | **-27.8** | **-36.9** | -26.7 | -8.8 | -3.2 | -11.9 |
| - | X | X | -22.4 | -25.6 | -36.4 | -25.1 | -3.2 | 0.8 | -11.0 |
| X | X | X | -17.9 | -22.4 | -32.3 | -21.5 | -5.4 | 0.8 | -6.3 |
| Spline models | | | | | | | | | |
| X | - | - | -15.1 | -17.1 | -24.6 | -19.9 | -8.1 | -4.8 | **-15.8** |
| - | X | - | -10.5 | -10.4 | -16.7 | -16.1 | 1.4 | -1.5 | -3.6 |
| - | - | X | -6.2 | -14.0 | -17.4 | -7.2 | -0.7 | 0.6 | 5.1 |
| X | X | - | -9.4 | -11.7 | -21.3 | -15.9 | -1.5 | **-6.5** | -12.7 |
| X | - | X | -18.4 | -24.2 | -32.9 | -18.4 | -3.4 | 0.7 | -8.7 |
| - | X | X | -22.1 | -25.6 | -36.4 | -25.1 | 3.8 | 5.0 | -5.4 |
| X | X | X | -12.0 | -18.5 | -27.9 | -12.2 | 13.6 | 9.3 | -8.0 |

Table S5. $R_t$ in Winter vs Summer, estimated from main analyses with datasets of different weekdays. Grey cells match those of Table S2.

| Model terms | | | Weekday | | | | | | |
|---|---|---|---|---|---|---|---|---|---|
| *T* | *ah* | *sr* | Fri | Sat | Sun | Mon | Tue | Wed | Thu |
| Linear models | | | | | | | | | |
| - | - | - | 1 (1;1) | 1 (1;1) | 1 (1;1) | 1 (1;1) | 1 (1;1) | 1 (1;1) | 1 (1;1) |
| X | - | - | 1.4 (1.2;1.7) | 1.4 (1.2;1.7) | 1.5 (1.3;1.7) | 1.4 (1.2;1.5) | 1.2 (1.1;1.4) | 1.2 (1.1;1.4) | 1.3 (1.1;1.5) |
| - | X | - | 1.4 (1.2;1.8) | 1.4 (1.1;1.7) | 1.5 (1.2;1.8) | 1.3 (1.2;1.6) | 1.2 (1.0;1.3) | 1.1 (1.0;1.3) | 1.3 (1.1;1.5) |
| - | - | X | 1.3 (1.1;1.6) | 1.3 (1.1;1.6) | 1.3 (1.2;1.5) | 1.3 (1.1;1.5) | 1.2 (1.0;1.4) | 1.2 (1.0;1.4) | 1.1 (1.0;1.3) |
| X | X | - | 1.4 (1.2;1.7) | 1.4 (1.2;1.7) | 1.4 (1.2;1.7) | 1.3 (1.2;1.5) | 1.2 (1.1;1.3) | 1.2 (1.1;1.3) | 1.3 (1.1;1.5) |
| X | - | X | 1.7 (1.4;2.1) | 1.6 (1.4;1.9) | 1.6 (1.4;1.9) | 1.5 (1.3;1.7) | 1.3 (1.1;1.6) | 1.3 (1.1;1.6) | 1.4 (1.2;1.6) |
| - | X | X | 1.9 (1.5;2.4) | 1.7 (1.4;2.2) | 1.8 (1.5;2.2) | 1.6 (1.3;1.9) | 1.4 (1.1;1.6) | 1.3 (1.1;1.6) | 1.5 (1.2;1.8) |
| X | X | X | 1.8 (1.4;2.3) | 1.7 (1.3;2.1) | 1.7 (1.4;2.2) | 1.5 (1.2;1.9) | 1.3 (1.1;1.5) | 1.3 (1.1;1.5) | 1.4 (1.2;1.8) |
| Spline models | | | | | | | | | |
| X | - | - | 1.8 (1.4;2.3) | 1.6 (1.3;1.9) | 1.6 (1.3;2.0) | 1.4 (1.2;1.8) | 1.3 (1.1;1.6) | 1.4 (1.2;1.8) | 1.6 (1.3;2.1) |
| - | X | - | 1.4 (1.2;1.8) | 1.4 (1.1;1.7) | 1.5 (1.2;1.9) | 1.3 (1.2;1.6) | 1.2 (1.1;1.4) | 1.2 (1.1;1.5) | 1.4 (1.1;1.7) |

| | | | | | | | | | |
|---|---|---|---|---|---|---|---|---|---|
| - | - | X | 1.3 (1.1;1.6) | 1.3 (1.1;1.6) | 1.3 (1.2;1.5) | 1.3 (1.1;1.5) | 1.2 (1.0;1.4) | 1.2 (1.0;1.4) | 1.2 (1.1;1.4) |
| X | X | - | 1.8 (1.4;2.3) | 1.5 (1.3;1.9) | 1.6 (1.3;2.0) | 1.4 (1.2;1.8) | 1.3 (1.2;1.6) | 1.5 (1.3;1.8) | 1.6 (1.3;2.1) |
| X | - | X | 2.0 (1.5;2.6) | 1.7 (1.4;2.1) | 1.7 (1.4;2.1) | 1.5 (1.2;1.9) | 1.3 (1.1;1.7) | 1.5 (1.2;1.9) | 1.8 (1.4;2.3) |
| - | X | X | 1.9 (1.5;2.4) | 1.8 (1.4;2.2) | 1.8 (1.5;2.2) | 1.6 (1.3;1.9) | 1.3 (1.1;1.6) | 1.4 (1.2;1.8) | 1.7 (1.3;2.2) |
| X | X | X | 2.0 (1.5;2.6) | 1.7 (1.4;2.2) | 1.8 (1.4;2.3) | 1.5 (1.2;1.9) | 1.3 (1.2;1.6) | 1.5 (1.2;1.9) | 1.7 (1.3;2.2) |

*Dataset with a single period between 26 June 2021 and 9 July 2021*

For the main analysis, the two-week period from 26 June to 9 July 2021 was split into two separate periods, because of the large influence of one of these observations as measured by Cook's distance (Supplementary Data, section 1).

If the period were treated as one, a more complex model would have been selected (solar radiation and temperature, or solar radiation and absolute humidity, with interactions), with estimated differences of $R_t$ in winter vs summer of 2.4 (95% CI: 1.8; 3.0) or 2.9 (2.1; 4.1), instead of 1.7 (1.4; 2.1) or 1.9 (1.5; 2.4) in the main analysis (Table S6). It shows that indeed this single period has a major influence on the results, supporting the decision to split the period into two one-week periods, which effectively removes these observations from estimating the weather effect.

Table S6. Results of the main analyses carried out with a dataset with a single two-week period between 26 June 2021 and 9 July 2021. The lowest $\Delta AIC_c$ (indicating highest support) is underlined.

| | Model coefficients (SE) [a] | | | | $R_t$ winter vs summer (95% CI) | Performance | |
|---|---|---|---|---|---|---|---|
| | Temperature | Absolute humidity | Solar radiation | Interaction [b] | | $\Delta AIC_c$ [c] | $R^2$ [d] |
| 0 | - | - | - | - | 1 | 0 | 0.41 |
| 1 | -0.026 (0.008) | - | - | - | 1.5 (1.2 ; 1.9) | -12.0 | 0.49 |
| 2 | - | -0.045 (0.021) | - | - | 1.4 (1.1 ; 1.9) | -1.6 | 0.44 |
| 3 | | | -0.26 (0.06) | - | 1.6 (1.3 ; 2.0) | -19.7 | 0.53 |
| 4 | -0.056 (0.017) | 0.088 (0.045) | - | - | 1.4 (1.2 ; 1.8) | -12.7 | 0.51 |
| 5 | -0.097 (0.020) | 0.132 (0.044) | - | -0.0073 (0.0022) | 2.1 (1.5 ; 3.0) | -24.4 | 0.58 |
| 6 | -0.021 (0.007) | - | -0.23 (0.06) | - | 2.0 (1.6 ; 2.7) | -26.9 | 0.57 |
| | **-0.029 (0.007)** | **-** | **-0.22 (0.06)** | **-0.029 (0.009)** | **2.4 (1.8 ; 3.0)** | **-39.3** | **0.63** |
| 7 | - | -0.049 (0.019) | -0.27 (0.06) | - | 2.2 (1.6 ; 3.0) | -24.7 | 0.56 |
| | **-** | **-0.068 (0.018)** | **-0.33 (0.06)** | **-0.067 (0.018)** | **2.9 (2.1 ; 4.1)** | **-39.2** | **0.63** |
| 8 | -0.023 (0.015) | 0.006 (0.050) | -0.22 (0.07) | - | 2.0 (1.5 ; 2.9) | -21.3 | 0.57 |
| | -0.025 (0.018) | -0.009 (0.046) | -0.27 (0.07) | -0.068 (0.018) | 2.6 (1.9 ; 3.8) | -36.7 | 0.64 |

*Analysis without offsets*

The variable *period* was primarily included to reflect changes in social distancing measures, but was also meant to absorb more gradual changes in SARS-CoV-2 transmission not explicitly included in the offsets or other variables. Whether that is properly done can be tested by repeating the main analysis without offsets, the effect of which should be taken over by *period*. It turns out that the model selection and estimated weather effects are hardly affected (Table S7), and that indeed the estimated *period* effects from the model without offset are almost identical to the sum of the period effects and offsets from the model with offset (Figure S3).

Table S7. Results of the main analysis, without the offsets. The model with lowest $\Delta AIC_c$ is indicated in bold.

| | Model coefficients (SE) [a] | | | | $R_t$ winter vs summer (95% CI) | Performance | |
|---|---|---|---|---|---|---|---|
| | Temperature | Absolute humidity | Solar radiation | Interaction [b] | | $\Delta AIC_c$ [c] | $R^2$ [d] |
| 0 | - | - | - | - | 1 | 0 | 0.68 |
| 1 | -0.021 (0.006) | - | - | - | 1.4 (1.2 ; 1.7) | -16.0 | 0.74 |
| 2 | - | -0.043 (0.015) | - | - | 1.4 (1.1 ; 1.7) | -6.9 | 0.71 |
| 3 | | | -0.19 (0.05) | - | 1.4 (1.2 ; 1.7) | -15.5 | 0.73 |
| 4 | -0.035 (0.013) | 0.039 (0.034) | - | - | 1.4 (1.2 ; 1.6) | -12.6 | 0.74 |
| 5 | -0.063 (0.016) | 0.073 (0.034) | - | -0.0048 (0.0017) | 1.8 (1.4 ; 2.3) | -19.9 | 0.76 |
| **6** | **-0.018 (0.005)** | **-** | **-0.16 (0.05)** | **-** | **1.7 (1.4 ; 2.1)** | **-27.2** | **0.77** |
| **7** | **-** | **-0.045 (0.014)** | **-0.19 (0.05)** | **-** | **1.9 (1.5 ; 2.4)** | **-26.6** | **0.77** |
| 8 | -0.011 (0.014) | -0.019 (0.037) | -0.17 (0.05) | - | 1.8 (1.4 ; 2.3) | -21.8 | 0.77 |

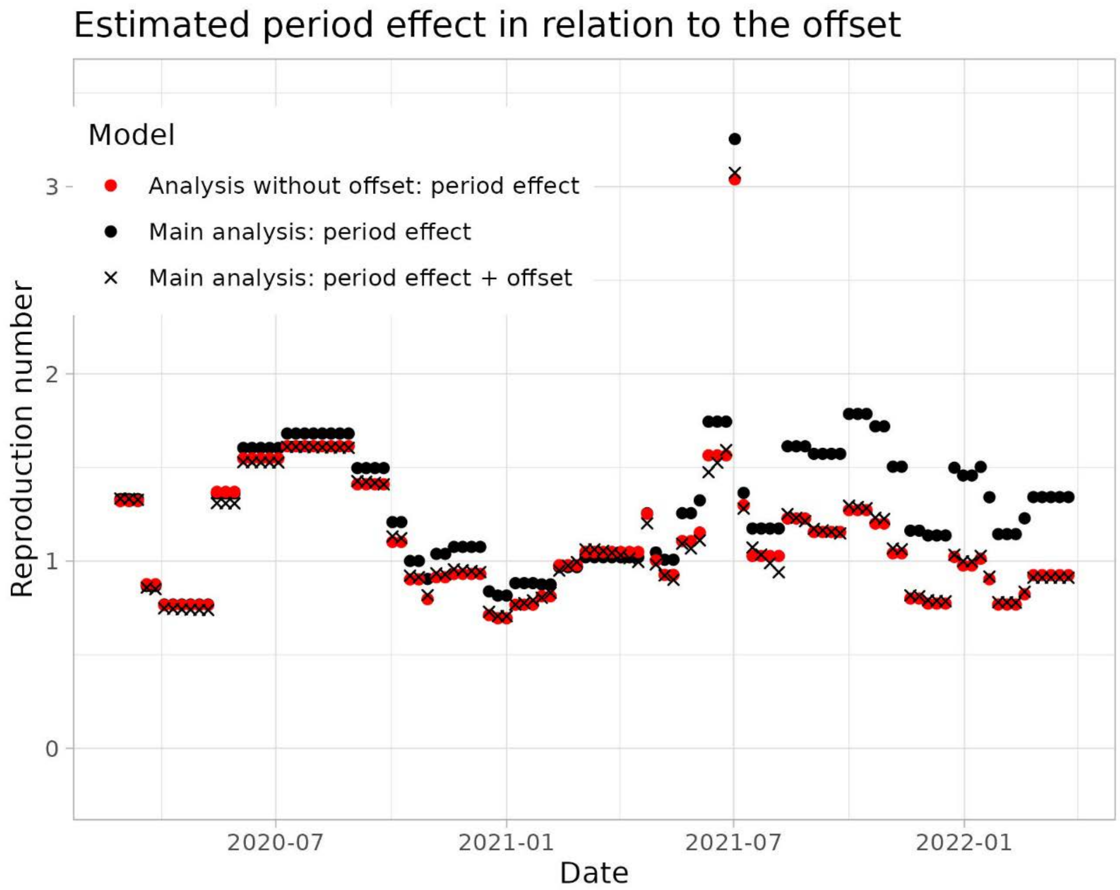


Figure S4. The estimated period effects of the best models (linear with solar radiation and temperature) without and with offset terms. The estimated period effect of the model without offset is almost identical to the sum of the offset and the period effects of the model of the main analysis (including offset).
